# Health factors associated with development and severity of post-stroke dysphagia: an epidemiological investigation

**DOI:** 10.1101/2023.08.29.23294807

**Authors:** Brittany N Krekeler, Heidi J P Schieve, Jane Khoury, Lili Ding, Mary Haverbusch, Kathleen Alwell, Opeolu Adeoye, Simona Ferioloi, Jason Mackey, Daniel Woo, Matthew Flaherty, Felipe De Los Rios La Rosa, Stacie Demel, Michael Star, Elisheva Coleman, Kyle Walsh, Sabreena Slavin, Adam Jasne, Eva Mistry, Dawn Kleindorfer, Brett Kissela

## Abstract

**Background and Purpose:** Dysphagia is a common post-stroke occurrence and has been shown to impact patients’ morbidity and mortality. The purpose of this study was to use a large population-based dataset to determine specific epidemiological and patient health risk factors that impact development and severity of dysphagia after acute stroke.

**Methods:** Using data from the Greater Cincinnati Northern Kentucky Stroke Study, GCNKSS, involving a representative sample of approximately 1.3 million people from Southwest Ohio and Northern Kentucky of adults (age ≥18), ischemic and hemorrhagic stroke cases from 2010 and 2015 were identified via chart review. Dysphagia status was determined based on bedside and clinical assessments, and severity by necessity for alternative access to nutrition via nasogastric (NG) or percutaneous endoscopic gastrostomy (PEG) tube placement. Comparisons between patients with and without dysphagia were made to determine differences in baseline characteristics and pre-morbid conditions. Multivariable logistic regression was used to determine factors associated with increased risk of developing dysphagia.

**Results:** Dysphagia status was ascertained from 4139 cases (1709 with dysphagia). Logistic regression showed: increased age, Black race, higher NIHSS score at admission, having a hemorrhagic stroke (vs infarct), and right hemispheric stroke increased risk of developing dysphagia after stroke. Factors associated with reduced risk included history of high cholesterol, lower pre-stroke mRS score, and white matter disease.

**Conclusions:** This study replicated many previous findings of variables associated with dysphagia (older age, worse stroke, right sided hemorrhagic lesions), while other variables identified were without clear biological rationale (e.g. Black race, history of high cholesterol and presence of white matter disease). These factors should be investigated in future, prospective studies to determine biological relevance and potential influence in stroke recovery.

## Introduction

The development of swallowing impairments (dysphagia) has been shown to have an impact on hospitalization timeline and comorbidities associated with acute stroke recovery. ^1^ Specifically, patients with dysphagia have poor nutrition and hydration, ^1^ both of which are critical for neural recovery after stroke. ^1,2^ Dysphagia is also associated with higher risk of pneumonia, a serious and deadly complication of swallowing impairment. ^3–10^

Prior investigations of dysphagia after ischemic or hemorrhagic stroke have focused primarily on characterizing specific impairments in swallow physiology based on lesion location and size, ^11–13^ or reporting on specific outcomes of these impairments in this population. ^3,14^ These investigations are highly relevant for assessing and treating the disorder and understanding disposition of these patients. However, patient-specific factors related to increased risk of developing dysphagia after stroke, and patient characteristics linked to severity of dysphagia, are not well understood. These patient-specific factors include both pre-morbid health conditions (e.g. heart disease) and patient demographics (e.g. sex, age, race), which in-part derive social determinants of health (e.g. economic stability and health literacy/education) ^15^ and are known to impact health outcomes at multiple levels. ^16^ Gaining an understanding of underlying conditions, biological, and socioeconomic risk factors that may predispose patients to developing swallowing disorders after stroke would provide opportunities for preventative care. To date, there have been no study populations large enough to address these gaps.

The goal of this work is to investigate incidence of dysphagia after stroke in a large retrospective, population-based dataset to determine what patient-specific factors may influence dysphagia status, or severity of impairment. These data will guide future investigations to better understand how and why certain individuals develop swallowing impairments after stroke and what can be done in acute clinical management of stroke to help prevent development of swallowing disorders.

## Methods

The Greater Cincinnati Northern Kentucky Stroke Study (GCNKSS) has been previously described in detail; ^17–21^ in brief, this is a population-based study using retrospective ascertainment of physician adjudicated stroke cases in a sample of approximately 1.3 million people across five counties in Southwestern Ohio and Northern Kentucky. All hospitalized adult (age ≥18) ischemic and hemorrhagic stroke cases from 2010 and 2015 were identified using International Classification of Diseases (ICD) stroke codes (ICD-9 430-436; ICD-10 I60-I69) at 15 area hospitals.

### Case Definition – Dysphagia vs No Dysphagia

Dysphagia status was identified through Electronic Medical Record (EMR) review by study team Registered Nurses. Individuals with documentation in the EMR of swallowing impairment were included in the “Dysphagia” group. Given the known variability in swallowing assessment practices both within and across institutions (e.g. timing of initial screening, formal evaluation by a speech-language pathologist vs bedside screen by nursing staff, etc), ^22,23^ documentation of dysphagia varied depending on the case, the treating institution, and clinical practices at time of stroke. Assessment of dysphagia status was also documented in the EMR. This could include a formal swallow assessment (bedside and/or instrumental by a speech-language pathologist) or a nursing bedside screen. Only individuals who were determined not to have dysphagia after one or more of these assessments were included in the “No Dysphagia” group. Within the identified “Dysphagia” group, individuals were compared based on their need for access to alternative nutrition during acute hospital stay. Individuals were categorized as one of the following (Dysphagia PO Subgroups): 1) Dysphagia not requiring PEG/NG (i.e. dysphagia management only required a modified texture and/or liquid diet), 2) Dysphagia requiring NG, 3) Dysphagia requiring PEG. PO status was determined and confirmed through same chart review procedures previously described. For cases where dysphagia status changed (i.e. progressed) throughout inpatient stay, the most invasive intervention required was documented (i.e. PEG was placed after NG).

### Patient Characteristics - Demographics, Clinical Presentation, Pre-Morbid Conditions

Patients included in the analyses were further characterized and compared (Dysphagia vs No Dysphagia, and Dysphagia PO Subgroups). The following patient characteristics were available for analysis, abstracted during the RN chart review process: 1) Patient demographic and social determinants of health data: age, sex, race, ethnicity, and estimation of a neighborhood Deprivation Index score; ^24^ 2) Clinical presentation at time of acute injury: National Institutes of Health Stroke Scale [NIHSS], pre-stroke Modified Rankin Scale [mRS], Glasgow Coma Score; 3) Stroke/lesion characteristics: type of case [ischemic/hemorrhagic], lesion location [hemisphere], ischemic stroke subtype [small vessel, cardioembolic, large vessel]; 4) Comorbidities (pre-morbid conditions): history of hypertension, diabetes, elevated cholesterol, coronary artery disease, myocardial infarction [MI], congestive heart failure [CHF], prior stroke, prior transient ischemic attack [TIA], dementia, brain injury or tumor, history of smoking, current smoker (within the past six months), alcohol use, heavy alcohol use, and any white matter disease [WMD, new and/or old]. All data regarding race and ethnicity categorization were abstracted directly from the electronic medical record (EMR); as such we are unable to definitively determine the source of the EMR reporting of race and ethnicity, whether it was self-identification or assigned by the admitting medical professional.

### Statistical Analyses

Study variables were summarized using descriptive statistics (mean and standard deviation or median and interquartile range [IQR] and/or range for continuous variables and frequencies and percentages for categorical variables) by study groups (Dysphagia status: Yes vs. Tested and No; Dysphagia subgroups: Dysphagia Not Requiring PEG/NG vs. Dysphagia Requiring NG vs. Dysphagia Requiring PEG; assessment: assessed vs. not assessed). Distribution was examined for all continuous variables. Associations between categorical variables and study groups were tested using either Chi-square or Fisher’s exact tests. Associations between continuous variables and study groups were tested using either two-sample t-tests, analysis of variance (ANOVA), Wilcoxon rank sum tests, or Kruskal-Wallis tests. Comparisons between the three Dysphagia subgroups were adjusted for multiple comparisons using Bonferroni correction for categorical variables, NIHSS and Glasgow Coma Score and by applying Dunnett’s test, following ANOVA, for age and deprivation index. Multivariable logistic regression analysis was carried out for Dysphagia status (Yes vs. Assessed and No) and dichotomized Dysphagia subgroups (Dysphagia requiring PEG or NG versus Dysphagia not requiring PEG or NG). P-values less or equal to 0.05 were declared as statistically significant. All analysis was carried out in SAS^®^ 9.4 (SAS Institute, Cary NC).

## Results

A total of 5592 (2010 N=2644; 2015 N=2948) physician adjudicated and confirmed hospitalized strokes from 15 area hospitals in the Greater Cincinnati and Northern Kentucky region were reviewed.

### Dysphagia Status and Dysphagia Sub-Group

Dysphagia status could only be ascertained from 4139 cases, which made up the final sample for the analyses (**Figure 1**). Out of 4139, 1709 cases were determined to have dysphagia. Of these cases, 944 did not require alternative access to nutrition (i.e. PEG/NG), 437 required NG, and 328 required PEG placement (**Table 1**). In comparing the subgroups by year, there were no differences between 2015 and 2010 in terms of proportion of individuals per Dysphagia Subgroup.

**Figure 1.**
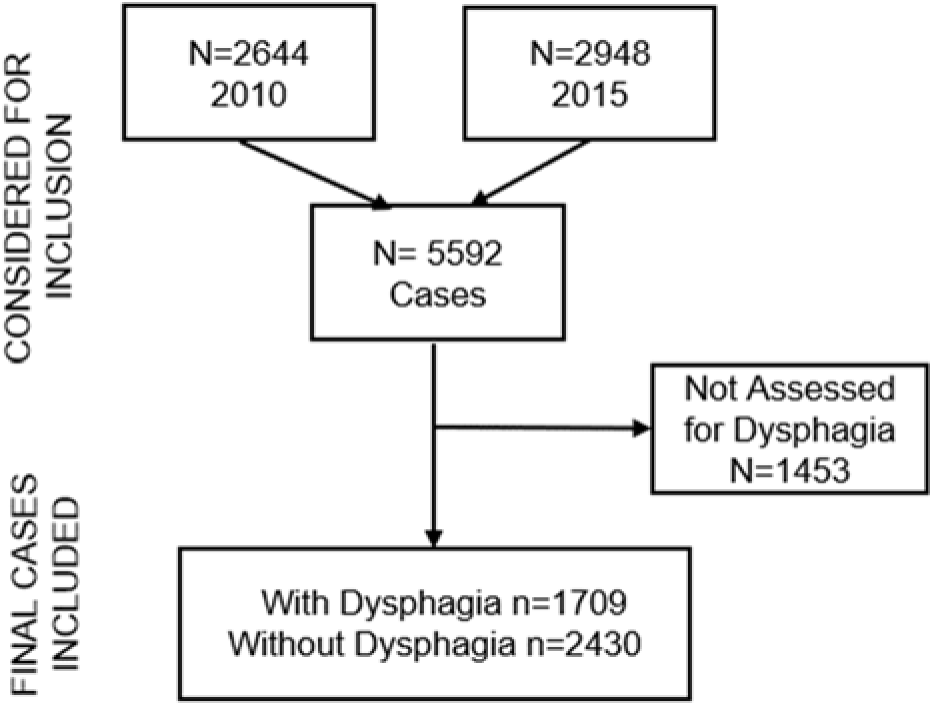
Number of Strokes included from Greater Cincinnati Northern Kentucky Stroke Study in 2010 and 2015.

**Table 1.** Dysphagia Subgroup Status by Year.

| **Dysphagia Subgroup**<br>n = 1709 | Dysphagia Not Requiring PEG/NG<br>n =944 | Dysphagia Requiring NG<br>n = 437 | Dysphagia Requiring PEG<br>n = 328 |
| --- | --- | --- | --- |
| **Year** |  |  |  |
| **2010** (n=782) | 406 (51.9%) | 223 (28.5%) | 153 (19.6%) |
| **2015** (n=927) | 538 (58.0%) | 214 (23.1%) | 175 (18.9%) |
| p-value<br>(2010 vs 2015) | 0.03 | 0.03 | 1.0 |

\*All p-values are Bonferroni adjusted

### Patient Characteristics – Demographics and Clinical Presentation

Patients with dysphagia were significantly older (mean 72 years vs 67.2 years p<0.0001), a larger proportion were females (57% vs 52.2%, p=0.002), had higher median NIHSS scores (8 vs 2; p<0.0001), lower median Glasgow Coma scores (14 vs 15, p<0.0001), higher median pre-stroke mRS scores (2 vs 1, p<0.0001), and scored slightly higher on the mean deprivation index (p=0.01) (**Table 2**).

**Table 2.** Demographics and Clinical Presentation by Dysphagia Status.

|  | Dysphagia | No Dysphagia | p-value |
| --- | --- | --- | --- |
| **N** | 1709 | 2430 |  |
| **Age, year, mean (std)** | 72.0 (14.7) | 67.2 (14.8) | <.0001 |
| **Race** |  |  | 0.06 |
| White + other | 1278 (74.8%) | 1877 (77.3%) |  |
| Black | 431 (25.2%) | 551 (22.7%) |  |
| Unknown* | 1 (0.06%) | 1 (0.04%) |  |
| **Ethnicity** |  |  | 0.02 |
| Hispanic | 6 (0.4%) | 14 (0.6%) |  |
| Non-Hispanic | 1211 (70.9%) | 1804 (74.3%) |  |
| Unknown | 492 (28.8%) | 611 (25.2%) |  |
| **Sex (M vs F)** |  |  | 0.002 |
| Males | 735 (43.0%) | 1161 (47.8%) |  |
| Females | 974 (57.0%) | 1268 (52.2%) |  |
| **Baseline NIHSS** |  |  | <.0001 |
| Median (25<sup>th</sup>, 75<sup>th</sup> %ile) | 8 (3, 16) | 2 (1, 5) |  |
| [min, max] | [0, 40] | [0, 39] |  |
| **Glasgow Coma Score** |  |  | <.0001 |
| Median (25<sup>th</sup>, 75<sup>th</sup> %ile) | 14 (11, 15) | 15 (15,15) |  |
| [min, max] | [3, 15] | [3, 15] |  |
| **Baseline mRS** |  |  | <.0001 |
| Median (25<sup>th</sup>, 75<sup>th</sup> %ile) | 2 (0, 3) | 1 (0, 3) |  |
| [min, max] | [0, 5] | [0, 5] |  |
| **Baseline mRS 0-1** | 610 (35.8%) | 1280 (52.7%) | <.0001 |
| **Deprivation Index, mean (std)** | 0.38 (0.13) | 0.37 (0.13) | 0.01 |

\*Unknown not included in statistical comparison

Stroke lesion characteristics between Dysphagia groups differed (**Table 3**): the Dysphagia group had a larger proportion of hemorrhagic strokes as compared to those with No Dysphagia (16.5% vs 11.8%, p<0.0001); the Dysphagia group had a greater proportion of “any right hemisphere” stroke when compared to the No Dysphagia group (51.3% vs 45.8%, p=0.002); amongst ischemic stroke cases, the Dysphagia group had a smaller proportion of small vessel strokes (9.3% vs 20.7%, p<0.0001), a greater proportion of cardioembolic strokes (38.5% vs 21.9, p<0.0001), and a smaller proportion of strokes where subtype could not be determined (32% vs 38.2%, p=0.0008).

**Table 3.** Lesion Characteristics by Dysphagia Status.

|  | Dysphagia | No Dysphagia | p-value<sup>345</sup> |
| --- | --- | --- | --- |
| **N** | 1709 | 2430 |  |
| **Type of Case** |  |  | <.0001* |
| Infarct | 1426 (83.4%) | 2142 (88.1%) |  |
| Hemorrhagic (ICH/SAH) | 281 (16.5%) | 287 (11.8%) |  |
| Unknown† | 2 (0.1%) | 1 (0.03%) |  |
| **Lesion Location(s)** |  |  | <.0001* |
| Left Hemisphere only | 681 (39.8%) | 1017 (41.8%) |  |
| Any Left Hemisphere | 946 (55.4%) | 1292 (53.2%) | 0.64** |
| Right Hemisphere only | 610 (35.7%) | 854 (35.1%) |  |
| Any Right Hemisphere | 877 (51.3%) | 1113 (45.8%) | 0.002** |
| Brainstem only | 4 (0.2%) | 7 (0.3%) |  |
| Any Brainstem | 153 (9.0%) | 187 (7.7%) | 0.57** |
| Mixed Hemispheric (L+R only) | 171 (10.0%) | 165 (6.8%) |  |
| Mixed (L and/or R + brainstem) | 149 (8.7%) | 180 (7.4%) |  |
| Unknown | 94 (5.5%) | 207 (8.5%) |  |
| **Stroke subtype Infarct only** |  |  | <.0001* |
| Small vessel (1) | 132 (9.3%) | 444 (20.7%) | <.0001** |
| Cardioembolic (2) | 549 (38.5%) | 470 (21.9%) | <.0001** |
| Large vessel (3) | 189 (13.3%) | 294 (13.7%) | 0.97** |
| Other identified (5) | 99 (7.0%) | 116 (5.4%) | 0.24** |
| Undetermined (8) | 456 (32.0%) | 818 (38.2%) | 0.0008** |

\*overall significance; \*\*after correcting for multiple testing; †unknown not included in the analysis

### Patient Characteristics - Pre-Morbid Conditions

A larger proportion of individuals in the Dysphagia group as compared to the No Dysphagia group had a history of hypertension (84.5% vs 81%, p=0.004), a history of coronary artery disease (36.2% vs 32%, p=0.0005), history of congestive heart failure (23.5% vs 15.6%, p<0.0001), history of prior stroke (28.4% vs 25.6%, p=0.04), and a history of dementia (16.7% vs 8.4%, p<0.0001) (**Table 4**). There were a larger proportion of individuals in the No Dysphagia group who had a history of TIA (16.6% vs 13%, p=0.002), who were current smokers (28.2% vs 23.8%, p=0.002) who reported alcohol use (at least 1-2 drinks per day; 41.7% vs 34.7%, p<0.0001), and those who had white matter disease (71.6% vs 68.5%, p=0.03).

**Table 4.** Pre-Morbid Conditions by Dysphagia Status.

|  | Dysphagia | No Dysphagia | p-value |
| --- | --- | --- | --- |
| **N** | 1709 | 2430 |  |
| History of Hypertension | 1444 (84.5%) | 1969 (81.0%) | 0.004 |
| History of Diabetes | 611 (35.8%) | 870 (35.8%) | 0.97 |
| History of Elevated Cholesterol | 944 (55.2%) | 1405 (57.8%) | 0.10 |
| Coronary Heart Disease | 794 (46.5%) | 938 (38.6%) | <.0001 |
| History of Coronary Artery Disease | 618 (36.2%) | 777 (32.0%) | 0.005 |
| History of Myocardial Infarction | 255 (14.9%) | 341 (14.0%) | 0.42 |
| History of Congestive Heart Failure | 401 (23.5%) | 379 (15.6%) | <.0001 |
| Prior Stroke | 486 (28.4%) | 621 (25.6%) | 0.04 |
| Prior TIA | 223 (13.0%) | 404 (16.6%) | 0.002 |
| History of Dementia | 286 (16.7%) | 204 (8.4%) | <.0001 |
| History of Brain Injury/Tumor | 12 (0.7%) | 15 (0.6%) | 0.84 |
| History of Smoking (Cigarettes) | 949 (55.5%) | 1400 (57.6%) | 0.18 |
| Current Smoker (Cigarettes) | 407 (23.8%) | 686 (28.2%) | 0.002 |
| Alcohol Use | 593 (34.7%) | 1013 (41.7%) | <.0001 |
| Heavy Alcohol use | 159 (9.3%) | 212 (8.7%) | 0.52 |
| White Matter Disease (any) | 1171 (68.5%) | 1739 (71.6%) | 0.03 |

### Logistic Regression Model for Factors related to Dysphagia Status

The variables that were considered for the logistic regression can be found in **Figure 2**. Variables that were included in the final model included age (per year increase), race (Black vs non-Black), baseline mRS score (0-1, i.e. functional vs 2-5), NIHSS (per point increase), Hemorrhage vs Infarct, lesion on the right side, high cholesterol, coronary artery disease, presence of white matter disease, deprivation index, and year of stroke (2015 vs 2010). The logistic regression model (ROC= 0.78; **Figure 2**) for dysphagia status revealed an increased risk of developing dysphagia after stroke for older individuals (Age/1 year OR=1.022, p<0.0001), people of Black race (OR=1.225, p=0.04), cases with higher NIHSS score (per point increase OR=1.133, p<0.001), cases with a hemorrhagic stroke vs an ischemic infarct (OR=1.472, p=0.0004), and having the stroke occur in the right hemisphere (OR=1.269, p=0.0011). Factors showing decreased risk of dysphagia included having lower baseline mRS scores of 0-1 (OR=0.657, p<0.0001), having high cholesterol (OR=0.84, p=0.02), having white matter disease (new or old, OR=0.801, p=0.01), and having been a case in 2015 (OR=0.82, p=0.007).

**Figure 2.**
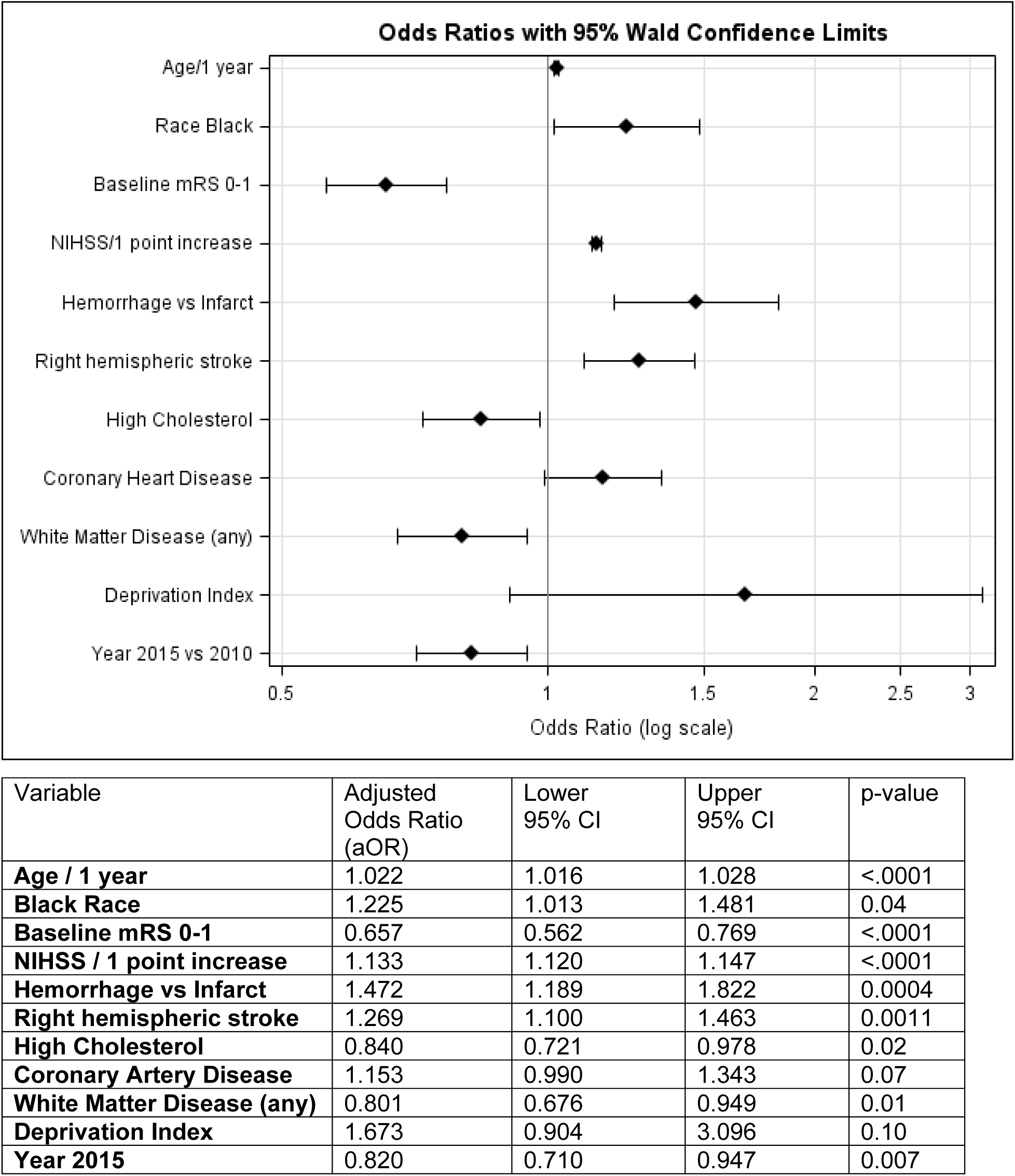
Logistic Regression by Dysphagia Status.

## Discussion

In this large, population-based study we sought to characterize differences in patient factors underlying dysphagia status for individuals after stroke and to determine which of these factors were associated with an increased risk in developing dysphagia post stroke.

Less than half of our final sample with positive dysphagia status required alternative access to nutrition via PEG or NG tube (**Table 1**). We found a greater proportion of individuals with dysphagia compared to those with no dysphagia were older, female, and had higher impairment scores upon stroke admission (i.e. NIHSS, GCS) and pre-stroke mRS (**Table 2**). It has been well established that increased age results in age-related changes to swallow movements and timing^25–29^ that may predispose older individuals to developing swallowing impairments after stroke. Regarding our finding associating female sex with swallowing impairments after stroke, a recent study showed that women had more severe outcomes after ischemic stroke as compared to men, which could result in worse swallowing outcomes after stroke. ^30^ However, another study of 578 patients in Seoul, South Korea found no relationship between swallowing outcomes after stroke and patient sex. ^31^ These alternative findings could be due to regional and cultural differences. Published evidence about influence of racial and ethnic background on stroke severity and outcome measures is mixed. ^32^ However, some literature suggests an ethnicity-age interaction, which could not be examined in these data, due to a small number of Hispanic individuals in our study population, could be influential. Other evidence points to some impact of socio-economic disparities amongst specific ethnic groups could be a contributing factor, ^33^ which is also supported by our data. Our data also showed that Deprivation Index scores were slightly higher in the Dysphagia group as compared to the No Dysphagia group, however, these were not found to be influential in the logistic regression (**Figure 2**). These discrepancies in the literature regarding the influence of sex and ethnicity in stroke-related outcomes, in general and specific to swallowing, highlight that more work is needed to investigate biological drivers for these differences.

Lesion type differed between the Dysphagia and No Dysphagia Groups, with a greater proportion of hemorrhagic, and right hemisphere strokes in the Dysphagia Group, which is supported by the literature. ^13,34,35^ Similarly, our findings of more dysphagia occurring with increasing stroke severity (NIHSS, Glasgow Coma score,), initial pre-stroke status as measured by mRS, hemorrhagic stroke, and right hemisphere lesions are also consistent with prior literature. ^13,35–40^ There have been some other studies, with smaller samples, ^41–43^ questioning the influence of laterality in development of dysphagia, but our findings provide more evidence towards hemispheric location of lesion being a potential risk factor for developing swallowing impairments. Our findings for stroke subtype also demonstrated that there were a greater proportion of cardioembolic strokes in the Dysphagia Group with ischemic stroke. This could be related to the increased likelihood of cardioembolic infarcts resulting in hemorrhagic transformation, ^44^ and as noted, hemorrhagic strokes may be more likely to present with dysphagia.

Regarding pre-morbid factors and development of dysphagia, we found that a greater proportion of individuals with a history of coronary artery disease, congestive heart failure, prior stroke, dementia, and hypertension in the Dysphagia Group as compared to the No Dysphagia Group. ^45,46^ Alcohol use, current cigarette smoking status, high cholesterol and prior TIA were also significantly different between the groups, with a higher proportion of individuals in the No Dysphagia group. While prior studies don’t directly study effects of alcohol consumption on development of dysphagia, alcohol use is known as risk factor for stroke. ^47,48^ However, studies have also shown that moderate alcohol use may reduce risk of ischemic stroke, ^49^ and that alcohol use does not affect stroke severity or outcomes after stroke. ^50^ These mixed results suggest that there may be a more complex relationship between alcohol consumption and stroke-related recovery, including swallowing, that has yet to be fully discovered.

Prior literature clearly demonstrate that individuals with cardiac conditions and dysphagia have a more difficult recovery and often poorer outcomes. ^51,52^ However, it is difficult to determine whether these cardiac conditions made patients more susceptible to developing dysphagia after stroke simply because these individuals are more medically compromised, or if there is a causal relationship between these conditions and swallowing impairments. In fact, our logistic regression results did not indicate increased risk of dysphagia development for patients with a history of coronary artery disease after accounting for other potential risk factors (**Table 4**), which may indicate the former hypothesis being more explanatory of these findings.

Other factors that were predictive of developing dysphagia after stroke included increased age, Black race, higher NIHSS score at presentation, having a hemorrhagic stroke (vs infarct), and right hemispheric stroke. Factors associated with decreased risk included patients having a history of high cholesterol, lower pre-stroke mRS, and any white matter disease detected on clinical imaging. Several findings were either surprising, or are still unclear in their meaning. As discussed, literature on ethnic and racial disparities in stroke severity and post-stroke care shows very mixed evidence and appears to be influenced by regional location and/or size of study. Prior published data have indicated that Black individuals have more severe strokes acutely than White individuals. ^53^ Current evidence provided in our study provides further emphasis of need for better understanding contributions of biological vs socioeconomic disparity to stroke-related outcomes in Black patients vs other racial categories.^54^

The findings regarding hyperlipidemia and any evidence of white matter disease on clinical stroke imaging being protective for risk of developing dysphagia after stroke were also unexpected. One hypothesis regarding the relationship between hyperlipidemia and lesser risk of dysphagia could be due to use of statin medications in this population. Literature supports the possibility that use of statins pre-stroke may reduce severity of stroke. ^55^ One study specifically looking at effects of statin therapy and development of post-stroke pneumonia suggested that patients who were using statins prior to admission for stroke and were treated with thrombolysis during admission were less likely to develop a pneumonia. ^56^ This finding should be explored more specifically in future studies of post-stroke dysphagia development, as this could be a potential protective treatment for individuals at risk of both stroke and dysphagia. In terms of any evidence of white matter disease serving as protective, this could be an incidental finding driven by interdependency among the many factors explored in this study. Given that we were unable to measure new white matter disease in these patients at the time of clinical imaging (i.e. differentiate new WMD from existing WMD before acute admission), it is difficult to tease out the meaning of this particular finding. Design of future studies of white matter disease and dysphagia could potentially include a cohort of repeat stroke patients with available prior clinical imaging in an attempt to better understand the relationship between existing and new WMD in post-stroke dysphagia.

## Limitations

The primary limitations of this study are due to the nature of retrospective analysis of data collected as part of a large data set; therefore, several broad and more nuanced factors were not available for use in our data analysis for both epidemiological factors and patient medical history. Most notably, this included the type of medical professional completing dysphagia evaluation and/or screening (e.g. speech-language pathologist or nursing professional). Further, the source of patient demographic factors (i.e. self-report vs determination by medical professional) related to race and ethnicity are not able to be determined in this type of retrospective dataset. These factors may have impacted classification of patients within certain groupings in our analysis. Further, the inability to differentiate new vs old WMD limits our ability to fully interpret the role of WMD in development of dysphagia after stroke.

## Conclusions

This large, population-based study confirms several factors that have been frequently demonstrated in the literature to be associated with developing dysphagia after stroke, including older age, dementia status, more severe stroke, and lesion type and location. However, other factors emerged that are not clear in their biological meaning or association, particularly findings related to certain racial/ethnic groups being significant factors for dysphagia development after stroke. These and other factors indicating a reduced risk, hyperlipidemia and any presence of white matter disease, should be considered in future prospective work to better understand these relationships and how these conditions may affect acute stroke recovery in the setting of dysphagia.

## Disclosures

BN Krekeler: Reports research support from the National Institutes of Health NIH/NCMRR/NICHD

HJP Schieve: No relevant disclosures

J Khoury: Reports research support from the National Institutes of Health NIH/NINDS

L Ding: No relevant disclosures

M Haverbusch: Reports research support from the National Institutes of Health NIH/NINDS K Alwell: No relevant disclosures

O Adeoye: Reports research support from the National Institutes of Health NIH/NINDS and Equity Holder and Co-Founder – Sense Diagnostics, Inc.

S Ferioloi: No relevant disclosures

J Mackey: Reports research support from the National Institutes of Health NIH/NINDS

D Woo: Reports research support from the National Institutes of Health NIH/NINDS

M Flaherty: Speaker’s bureau-CSL Behring and Alexion, Co-founder, employee, and equity holder: Sense Diagnostics, Inc., Clinical trial endpoint adjudication committee member: Boehringer Ingelheim and LG Chem

F De Los Rios La Rosa:

S Demel: No relevant disclosures

M Star: No relevant disclosures

E Coleman: No relevant disclosures

K Walsh: AstraZeneca: Speaker’s Bureau American Heart Association, NIH, Sense Diagnostics LLC, Jan Medical Inc: Grant Funding/Research Support

S Slavin: No relevant disclosures

A Jasne: No relevant disclosures

E Mistry: Reports research support from the National Institutes of Health NIH/NINDS

D Kleindorfer: No relevant disclosures

B Kissela: Reports research support from the National Institutes of Health NIH/NINDS

## Data Availability

All data are available to public upon request, please visit: https://www.gcnkss.com/ for details.

